# Effect of human mobility in Dengue spreading: Study cases for Caldas (CO)

**DOI:** 10.1101/2023.01.10.23284416

**Authors:** Carolina Ospina-Aguirre, David Soriano, Gerard Olivar-Tost, Cristian. C. Galindo-González, Jesús Gómez-Gardeñes, Gustavo Osorio

## Abstract

According to the World Health Organization (WHO), dengue is the most common acute arthropod-borne viral infection in the world. The spread of dengue and other infectious diseases is closely related to human activity and mobility. In this paper we analyze the effect on the total number of dengue cases within a population after introducing mobility restrictions as a public health policy. To perform the analysis, we use a complex metapopulation in which we implement a compartmental propagation model coupled with the mobility of individuals between the patches. This model is used to investigate the spread of dengue in the municipalities of Caldas (CO). Two scenarios corresponding to different types of mobility restrictions are applied. In the first scenario, the effect of restricting mobility is analyzed in three different ways: a) limiting the access to the endemic node but allowing the movement of its inhabitants, b) restricting the diaspora of the inhabitants of the endemic node but allowing the access of outsiders, and c) a total isolation of the inhabitants of the endemic node. In this scenario, the best simulation results are obtained when endemic nodes are isolated during a dengue outbreak, obtaining a reduction of up to 22.51% of dengue cases. Finally, the second scenario simulates a total isolation of the network, *i.e*., mobility between nodes is completely limited. We have found that this control measure reduces the number of total dengue cases in the network by up to 42.67%.

**Author summary:** For the World Health Organization, dengue is a disease of public health concern. In recent years there is an increasing trend in the number of dengue cases despite existing prevention and control campaigns. The mobility of the population is considered an important factor in dengue dispersion. In this paper, we are interested in addressing how restrictions to human mobility might affect the incidence of dengue in a region. Our research is relevant because the model can be adapted to other regions or scales, and the mobility control measures can be taken into account for the development of public health policies in endemic regions.

## 1 Introduction

According to the World Health Organization (WHO), dengue fever is the acute arthropod-borne viral infection with the highest incidence in humans in the world [1]. It is transmitted to humans mainly by the bite of a mosquito *Aedes aegypti* infected with Dengue virus (DENV). As a vector-borne disease the transmission between humans only occurs through the bite of infected mosquitoes, never from one person to another. In other words, the mosquito does not cause the disease directly, but acts as a bridge between two people, one with the virus and the other without it. In turn, the mosquito becomes infected when it feeds on the blood of a person infected with dengue and transmits the virus when it bites healthy people.

Mathematical models are extremely useful to understand mechanisms that drive a healthy population toward an epidemic or endemic state, as well as for evaluating containment measures that help to suppress, or at least mitigate, the incidence of a given communicable disease. The usual mathematical approach to such tasks is the use of compartmental models. In these models, individuals in a population can be divided into classes or compartments according to their epidemiological state. For instance, in the celebrated Susceptible-Infectious-Recovered (SIR) model, individuals are divided into susceptible (healthy people who may acquire the virus), infectious (people who have acquired and can transmit the virus) and recovered (people who cannot propagate the pathogen and have acquired immunity to the virus) [2–4]. Yet simple and minimal, the SIR model has been pervasively used to analyse a plethora of viral infections such as measles [5, 6], rubella [7], malaria [8], zika [9], COVID-19 [10, 11], dengue [12–18], among others.

In the case of vector-borne disease, more refined compartmental models have been introduced in which both human and vector populations are divided into several compartments. For instance in [19, 20] dynamics of human population is modeled through a SIR model whereas vectors are divided into Susceptible and Infectious. This division creates a SIR-SI model which is the compartmental dynamics adopted in our work. Alternative approaches for the study of the transmission of DENV include the Ross-Macdonald epidemic model [21] or the addition of further compartments to capture the growth dynamics of the vector population [22, 23]. In all these approaches, the main goal is to define control strategies that favor the eradication of the virus in humans population in a population.

Most of control strategies rely on improving hygiene measures, the use of pesticides or, more recently, the release of Wolbachia-infected vectors in high incidence habitats. However, human behavior plays an important role in the geographical spread of pathogens [24, 25], and vector-borne diseases are not the exception. One of the most salient features of human behavior affecting the spread of vector-borne diseases is human mobility [26]. For the particular case of dengue transmission, mobility determines the degree of exposure to the disease vectors [27, 28] and, evidently, it is essential to foster local dengue outbreaks in low incidence areas through the importation of cases from distant and high incidence regions [29, 30]. A remarkable recent example of such phenomenon was the reintroduction of the dengue virus in Singapore [31]. The contribution of human mobility in the spread of dengue cases has become an essential factor given the expansion of urban environments and the increased frequency of international travels [32–34]. It has been projected that by 2030 more than half of the world’s population will reside in urban areas in the tropics, due to population growth and migration from rural areas [35].

The importance of human behavior and, in particular, human mobility in the spread of communicable diseases has motivated the formulation of different mathematical frameworks to study the contribution of mobility in the spread of vector-borne diseases [12, 36]. Most of these approaches bridge the gap between single-population models to metapopulations by incorporating the complex architecture of human flows in the form of networks [37] defined through data-driven frameworks [38–40].

Equipped with this framework, we focus on the analysis of how mobility constraints affect dengue transmission considering two control scenarios. In the first, mobility constraints are concentrated in the nodes with the highest incidence. In scenario, the entire network is connected and two types of mobility restrictions are analyzed.

The first measure is to allow residents to leave but prevent outsiders from entering. In this scenario, preventing outsiders from entering and allowing residents to leave a high-incidence municipality was found to be the most effective measure, resulting in a 22.51% reduction in the number of dengue cases in the entire region. The second is to allow outsiders to enter a high-incidence municipality, but not to allow its residents to leave. And finally isolation consists of preventing residents from leaving and outsiders from entering a municipality. The second scenario analyzes the effect of confinement, *i.e*., totally restricting mobility throughout the network, preventing visitors from entering and residents from leaving a municipality. When isolation is applied, a reduction of cases of 42.67% is obtained compared to the unrestricted case.

This article is organized as follows. Section 2, Materials and methods, describes the proposed compartmental model, the methodology used for estimating the model parameters and the coupling of the epidemiological model with the mobility network. Section 3 presents the results of the case study in which the model was applied in the department of Caldas - Colombia, where we analyze the effectiveness of the different mobility restrictions described above. The manuscript rounds off in section 4 with a brief discussion of the results and their implication for informing public health decision makers.

## 2 Materials and methods

According to Satorras [37], we are currently witnessing a golden age in epidemic modeling: models are improving significantly thanks to the continuous addition of data, while the powerful computational resources available now make it possible to extend simulations to new limits. Throughout the years, SIR models have been used to analyse viral infections such as measles [5, 6], rubella [7], malaria [8], zika [9], COVID-19 [10, 11], dengue [12–18] and others.

### 2.1 SIR-SI compartmental model

Let us denote the total size of the populations for humans and mosquitoes by *N*_*h*_ and *N*_*m*_, respectively. Human population is divided into susceptible (*S*_*h*_), infectious (*I*_*h*_) and recovered (*R*_*h*_), while mosquitoes population is divided into susceptible (*S*_*m*_) and infectious (*I*_*m*_) ones. To define the model, the following assumptions are made:

- The size of the human population *N*_*h*_ is assumed to be constant, corresponding to the steady state on the underlying population dynamics, since the period of the disease is very small in relation with population dynamics.
- Incubation period is neglected for both, humans and mosquitoes.
- Deaths caused by the disease are not considered for either humans or mosquitoes.
- Mosquitoes cannot recover after being infected.
- There is not super-infection for either humans or mosquitoes.
- Susceptible humans can only get infected trough an infected mosquito bite.
- Human recruitment rate is constant.
- Mosquito recruitment rate depends on environmental conditions and is constant in each region.

The dynamics of dengue transmission between humans and mosquitoes, is defined by the system of equations (1). A susceptible mosquito can become infectious if it has contact with an infectious individual, according to the transmission probability of the vector *λ*_*m*_, the bite rate *β* and the proportion of infectious humans in the node 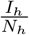. Therefore, the infection rate for susceptible mosquitoes is given by 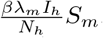. For the case of humans, the rate of infection is proportional to the biting rate *β* and the number of infectious mosquitoes *I*_*M*_, transmission probability *λ*_*h*_ and inversely proportional to the total population *N*_*H*_, determining the probability of a given individual being bitten. Therefore, the infection rate for susceptible humans is then expressed as 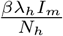.

Considering the former mechanisms, the dynamical evolution of each compartment associated to a given municipality is described by the following equations:

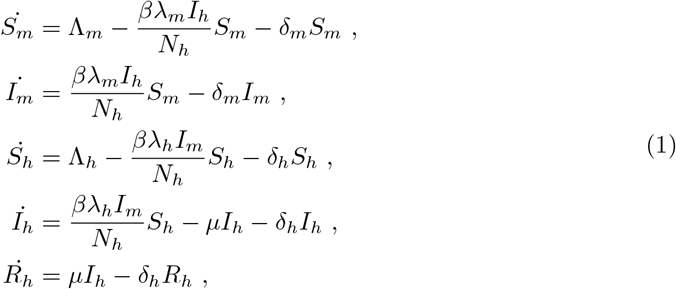

with *N*_*h*_ = *S*_*h*_ + *I*_*h*_ + *R*_*h*_. In table 1 we specify the definition of the parameters used in the former equations of the SIR-SI model.

**Table 1.** Parameters description for the SIR-SI model.

| Parameter | Description |
| --- | --- |
| $\beta$ | Average bite rate per time unit of the mosquito. |
| $\lambda_h$ | Probability of a human to become infectious. |
| $\lambda_m$ | Probability of a mosquito to become infectious. |
| $\delta_m$ | Natural death rate for mosquitoes. |
| $\mu$ | Recovery rate for humans. |
| $\delta_h$ | Natural death rate for humans. |
| $\Lambda_m$ | Recruitment rate of susceptible mosquitoes. |
| $\Lambda_h$ | Recruitment rate of susceptible humans. |

The dynamics of the infectious set in the SIR model according to *Allen* [41] can yield two different behaviors, one in which there is an epidemic peak and the other in which the number of infected individuals decreases monotonically until the end of the outbreak. In [42] the analysis developed by *Allen* is extended to the SIR-SI model (1) and it is shown that the dynamics for the infected population depends on

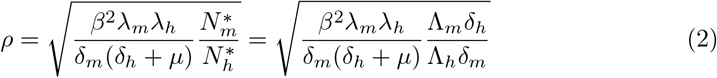

When *ρ >* 1, a maximum occurs (epidemic peak) and then decreases to zero. When these conditions generate *ρ <* 1 the infected have a decreasing behavior until they reach zero.

#### 2.1.1 Parameters estimation and initialization

The parameters governing contagion dynamics and the mosquito’s life-cycle are extracted from the work published by Helmersson *et al*. [43]. In this manuscript, the authors proposed different equations capturing the reduction of vectorial capacity of *Aedes aegypti* when being exposed to either very high (*T >* 34°*C*) or very low (*T <* 12°*C*) temperatures. The behavior of the parameters as a function of temperature can be seen in the Fig 1. In particular, the average bite rate reads:

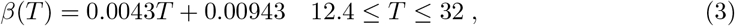

the probability that a mosquito becomes infectious after biting and infectious human is:

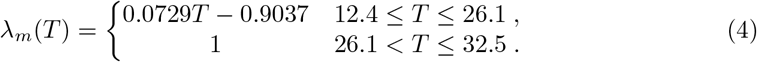

instead, in the range 12.286 ≤ *T* ≤ 32.461, the infection probability for humans can be modeled as:

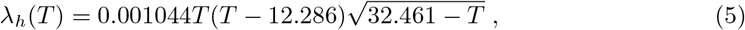

while the mosquito death rate goes as:

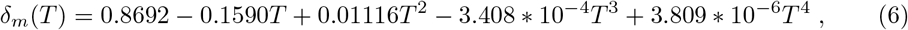

for 10.54 ≤ *T* ≤ 33.41. In Fig. 1 shows the variation of these four parameters as the temperature increases from 8°*C* to 40.7°*C*

**Fig 1.**
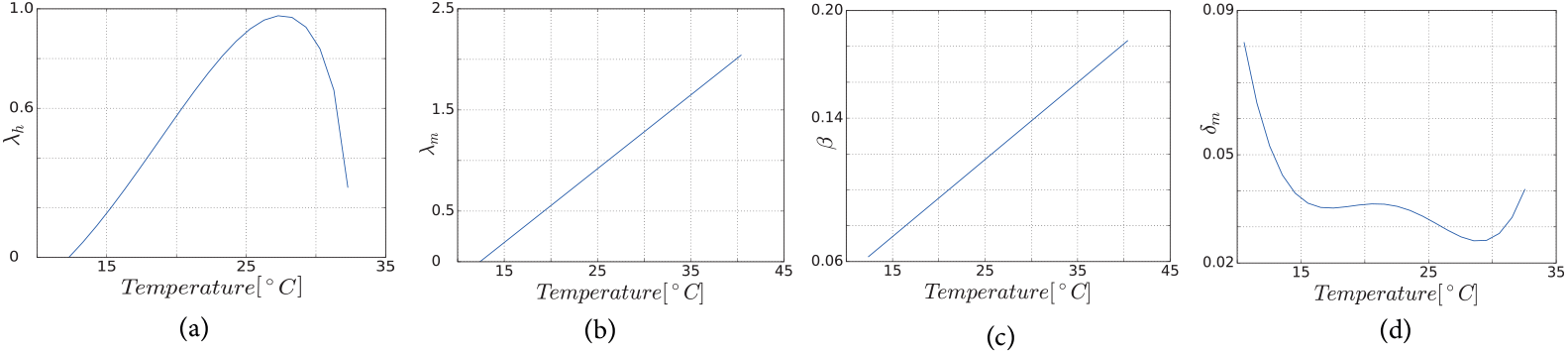
Temperature dependence of the parameters. (a) Bite rate *β*, (b) Rate of death of mosquitoes *δ*_*m*_, (c) Likelihood of a person becoming infectious *λ*_*h*_, (d) Likelihood of a mosquito becoming infectious *λ*_*m*_.

The other four parameters are defined as follows. For Λ_*h*_, the approach proposed by [44] is used, where it is defined as 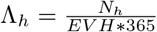. The recovery rate for humans *μ* is set to 0.32288 according to the study developed by Hamdan [14], while *δ*_*h*_ is estimated using known specific statistics from the geographical region under analysis. Finally, to estimate Λ_*m*_ we start from the equation for the vital dynamics of the vector when *S*_*m*_ = 0, (*i.e*. 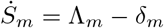), where the carrying capacity (number of mosquitoes at steady state) is obtained as 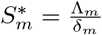. The recruitment rate, therefore, can be expressed in terms of the carrying capacity 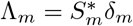.

To set the initial conditions for the model, some considerations have to be mentioned. First of all, it has to be noted that *N*_*h*_ can be fixed based on the known population of each municipality, while *N*_*m*_ can be estimated with the Index of Domiciliary Infestation (number of houses with one or more containers positive for immature *Aedes aegypti* divided by the number of houses sampled multiplied by 100) and the average number of people per household in the study region.

Considering Eqs. (1) and the parametrization described above we shown in Fig 2 the behavior of the model when simulating a region with an average temperature of 31°*C* (*β* = 0.227 *λ*_*m*_ = 1, *δ*_*m*_ = 0.0298, *λ*_*h*_ = 0.732, *μ* = 0.329) and the mosquito recruitment rate Λ_*m*_ = 86.634. When the initial conditions are *N*_*m*_ = 148.352, *S*_*m*_ = 118.352, *I*_*m*_ = 30, *N*_*h*_ = 76963 *S*_*h*_ = 76963 *I*_*h*_ = 0, that produces a *ρ >* 1 and an epidemic peak. When the initial conditions are *N*_*m*_ = 148.352, *S*_*m*_ = 118.352, *I*_*m*_ = 0, *N*_*h*_ = 76963, *S*_*h*_ = 76928, *I*_*h*_ = 35, that results in a *ρ <* 1 and thus the number of infected decreases to zero.

**Fig 2.**
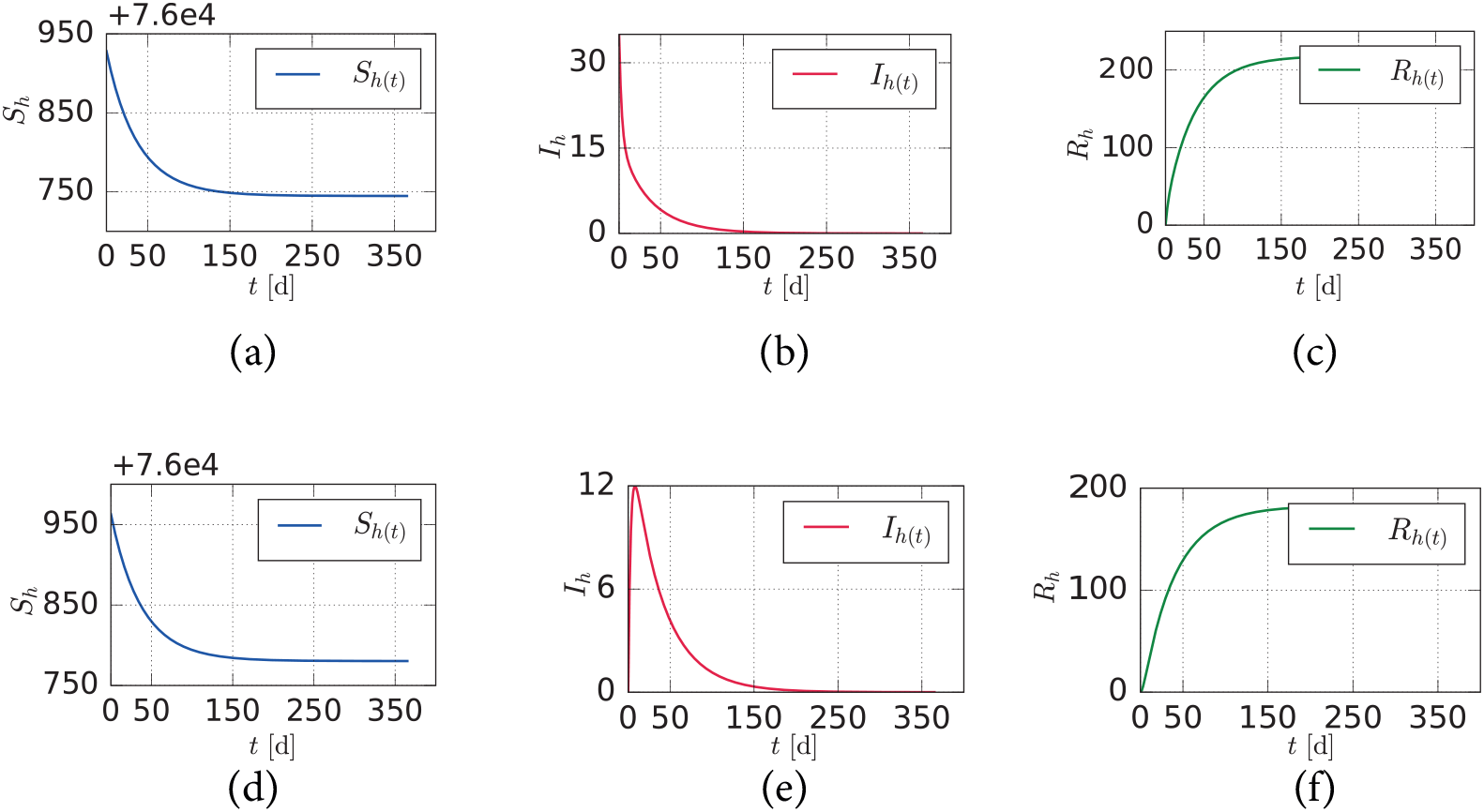
Behavior of the human population over time in the SIR-SI model. (a,c) Susceptible, (b,e) Infected and (c,f) Recovered. When *ρ <* 1 (a, b and c), and *ρ >* 1 (d,e and f).

### 2.2 SIR-SI model with human mobility

The SIR-SI can be modified to account for the impact of recurrent human mobility patterns. For this purpose, let us assume that each individual has his/her residence located inside a given patch but can move to other destination, identified as the workplace for instance. In the absence of data, we assume these movements to be governed by a fully-connected weighted matrix ϒ. We construct this matrix synthetically by assuming that 90% of the population inside each patch remain there whereas 10% of the population move to another area. We compute the flows connecting different patches following the gravity model [39], implying that the number of connections between two patches *i* and *j*, hereinafter denoted by *W*_*ij*_, can be expressed as:

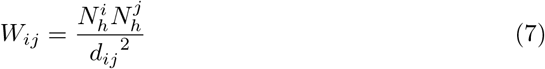

Taking into account the flows distribution, the elements of the mobility matrix ϒ read:

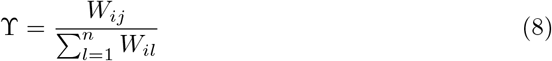

Agents’ movements between patches *i* and *j* cause a redistribution of the population across the system; therefore it is necessary to adjust the equations (1) to account for the actual number of people that are in any patch *i* at any given time. In particular, the effective population of patch *i* is defined as:

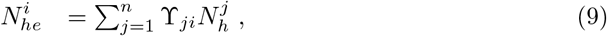

which accounts for the distribution of the residential population 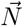 and the mobility patterns of the individuals of the metapopulation ϒ. Likewise, mobility also changes the effective number of infected individuals in each patch *i*, which now reads:

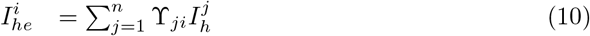

Finally, we assume that mosquitoes stay inside their associated area. To model the spatio-temporal evolution of the disease, we define the quantity *x*^*i*^(*t*) as the occupation of each compartment (*x* ∈ {*S*_*h*_, *I*_*h*_, *R*_*h*_, *S*_*m*_, *I*_*m*_}) inside each patch *i*. Following the assumptions of the model, the time evolution of these quantities is given by:

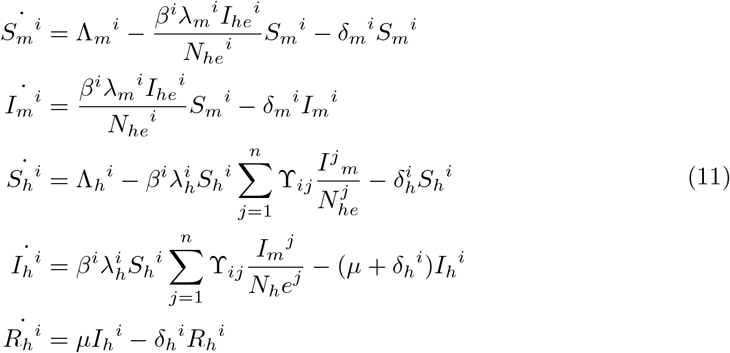

In Fig. 3 we show an scheme of the metapopulation model in which the SIR-SI compartmental model is integrated with human mobility flows.

**Fig 3.**
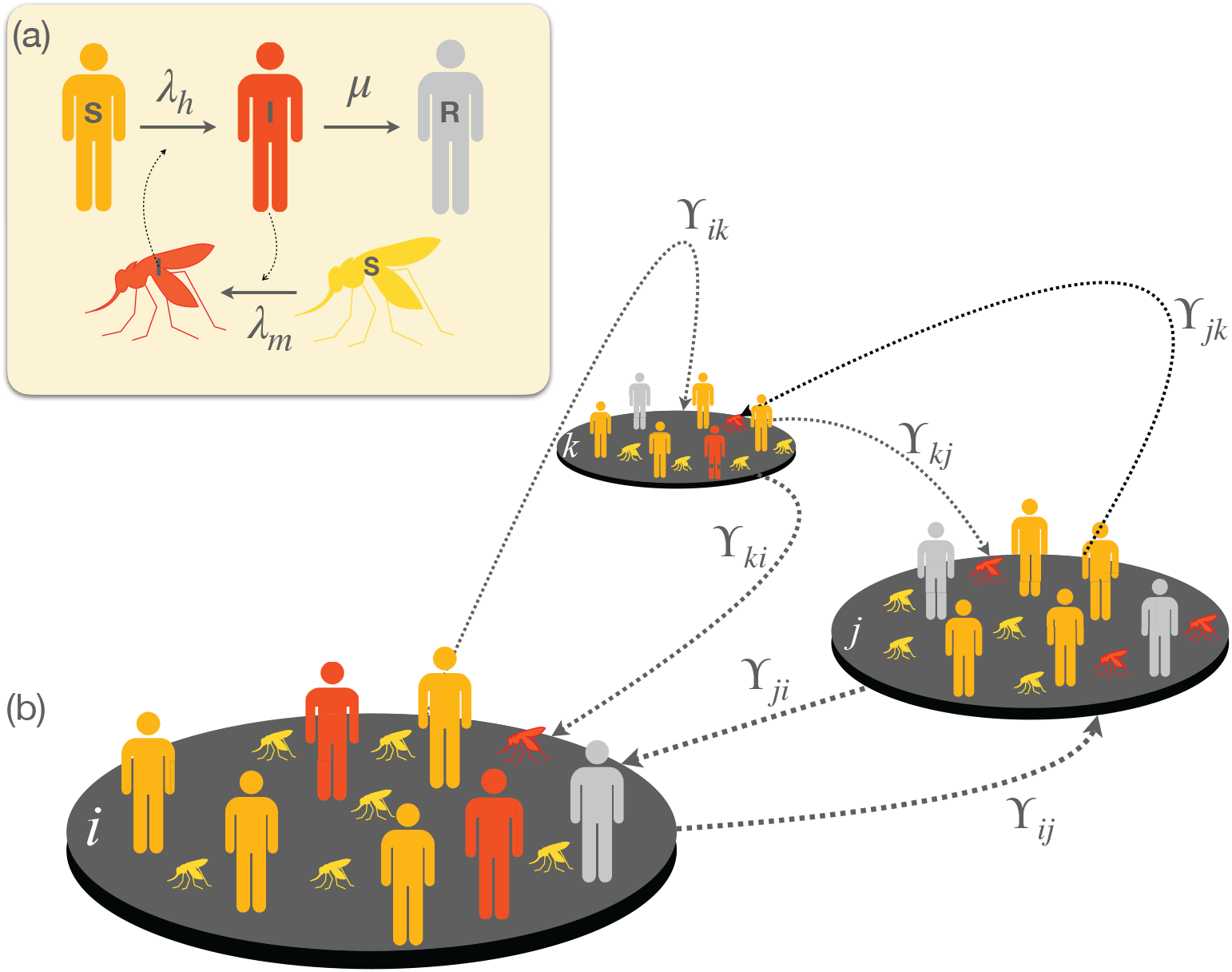
Schematic view of the SIR-SI metapopulation model. (a) shows the compartmental dynamics of the SIR-SI model with cross-infections between vectors and humans while panel (b) shows a toy metapopulation of 3 patches connected through links whose weights are given by matrix ϒ.

## 3 Results

Once presented the general formalism to tackle the dissemination of dengue fever across a metapopulation we now apply it to particular scenarios. We first address the analysis of three simple cases that illustrate how mobility between a small set of patches have profound implications in the overall extent of dengue cases. Then, we present our main case study: the spread of dengue fever in the department of Caldas (CO). We describe the main attributes of the department, their map into the metapopulation framework, and the parametrization of the model according to observed dengue cases. Finally, this section presents the results of two different mobility restriction and show their impact on the total dengue fever cases.

### 3.1 Mobility effects over dengue cases in simple metapopulations

We start by developing the analysis of three very simple metapopulations to unveil the effect of mobility on the spread of dengue. To this aim, we define three case examples in which the distance between nodes is set as a control parameter so that its increase implies, according to the gravity model, the decrease of mobility. The three scenarios are sketched in Fig. 4.

**Fig 4.**
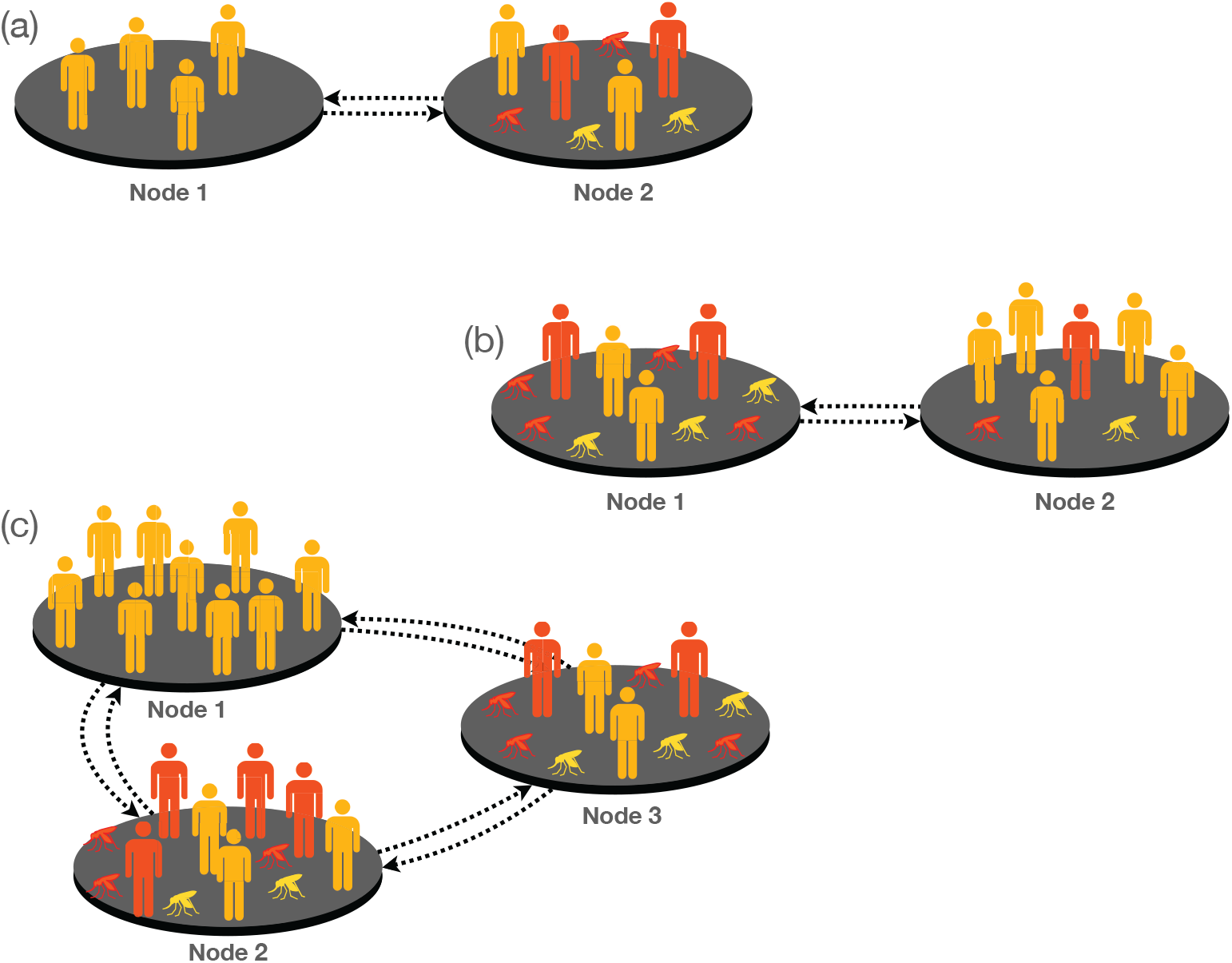
Schematic plot of the three simple metapopulations covered as case examples. In panel (a), corresponding to case A, node 1 is a zero-incidence node, i.e., there are no mosquitoes on it, however in node 2, there are environmental conditions for vector breeding and the disease is present. Panel (b) corresponds to case B. In this case node 1 is defined as endemic and is connected to a non-endemic node. Note that in both nodes the disease is present, but in node 1 the vector breeding conditions are better, so there is a wider spread of the disease than in node 2. Finally in panel (c) we define case C for which there are three equidistant connected nodes. Node 1 has zero incidence and its population is the largest while nodes 2 and 3 have characteristics of endemic regions. However, node 2 has more inhabitants than node 3 and, therefore, the probability of infection in node 3 is larger than in the other two patches.

#### 3.1.1 Case A

As sketched in Fig. 4.a this case example corresponds to a patch (node 1) with zero incidence connected to an endemic patch (node 2). The model parameters used to simulate this example are shown in Table 10(a). In Fig 5a, we show the evolution of dengue cases in both patches as the distance increases. This graph shows that increasing the distance between patches, i.e. reducing mobility between patches, leads to an overall reduction in the total number of cases. Note however that the effects of increasing distance (reducing mobility) for both patches are opposite. In particular, while increasing mobility increase the exposure of individuals from non-endemic areas leading to a higher incidence, movement across patches is beneficial for the population in endemic areas, decreasing the total incidence of the virus on this population.

**Fig 5.**
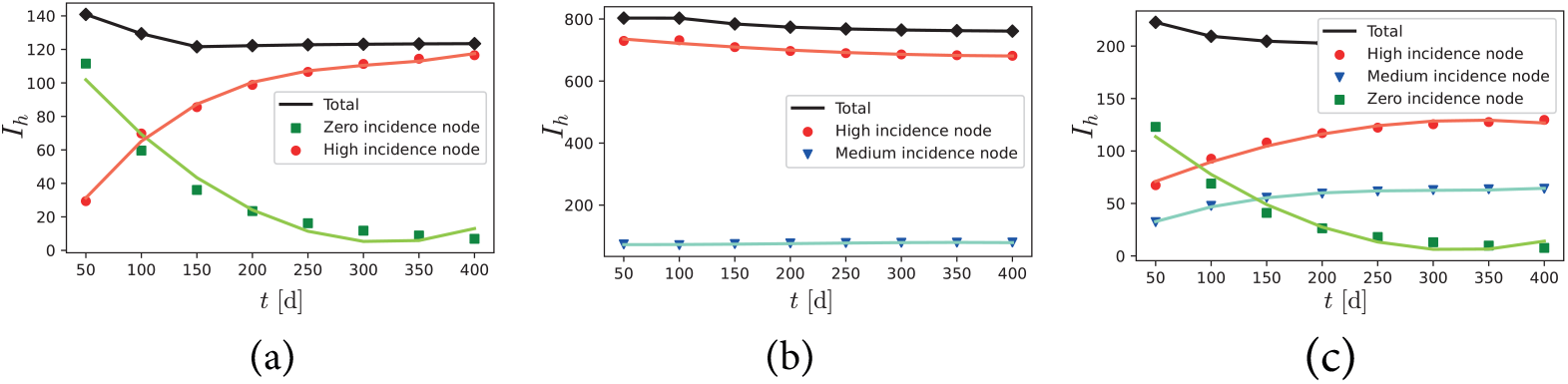
Evolution of dengue cases in patches as the distance increases when connecting a) High and a zero incidence node. b) High and medium incidence node. c) High, medium and zero incidence node. The green line shows the denge cases in the zero incidence node, the blue line shows the cases in the medium incidence node and the red line shows the cases in the high incidence node. The black line is used for the total number of cases.

#### 3.12 Case B

In this example, node 1 is a patch with high disease presence and node 2 a patch where there are fewer dengue cases (see Table 11(a) for the parameters used in this case). In Fig. 5b, we observe a qualitatively similar pattern to the one described before but much less pronounced due to the higher similarity among the connected patches.

#### 3.1.3 Case C

In this example three equidistant nodes are connected. According to the parameters chosen in Table 12, node 1 has zero incidence and the largest population, node 2 has a larger population than node 3, but the latter has a higher incidence of the disease (97.5 cases per thousand inhabitants versus 75.2 for node 2) when patches are isolated.

The results of this case are shown in Fig. 5 Case C. Again geographical location, *i.e*. the increase of the distance between patches, allows for a reduction in the total number of cases. However, locally,the situation of node 3 (zero incidence) worsens when the distance is reduced, while for the medium and high incidence nodes, proximity reduces the number of dengue cases by up to 50%.

The results of the former cases clearly show that mobility restrictions have a great impact on reducing the number of cases, although locally the epidemiological situation can worsen as a byproduct of this contention measures. Motivated by these results we now propose scenarios to evaluate the effect of applying mobility restrictions in a more complex metapopulation inspired in the municipalities of department of Caldas (CO).

### 3.2 Defining the metapopulation of the department of Caldas

The department of Caldas is located in the central west of the Colombian Andean region. It is crossed by the Central and Western mountain ranges, which makes it a department of mountainous topography. It has an area of 7888 *km*^2^ and a population of 987991 inhabitants [45], while its elevation above sea level ranges between 5400 masl and 170 masl. The department has 27 municipalities, ten of which have direct communication with 6 other municipalities in neighboring departments. To apply our SIR-SI model to the department of Caldas, we define a metapopulation with 33 patches, each with a number of residents equal to that reported by the census. The connections (links) between patches are calculated as a function of the road distance between them, resulting in the ϒ matrix containing the transition rates between pairs of municipalities. This matrix does not take into consideration whether the municipalities belong to the department of Caldas or they are external municipalities that by proximity have a relationship with the previous ones. The precise transition rates between the 33 municipalities (patches) are reported in Table 5).

For the numerical simulations, the parameters *β, λ*_*h*_, *λ*_*m*_ and *δ*_*m*_ are calculated with the procedure mentioned in section 2.1.1 and shown in Table 3 for each of the analyzed municipalities. The estimations are based on the average temperature value in each of them (see Table 2). In turn, the recruitment rate for humans is calculated with the value of life expectancy (*EV H* = 74.64), defined in public health records whereas humans mortality rate is obtained from DANE statistics.

**Table 2.** Average altitude and temperature of the municipalities of the department of Caldas. Source: Caldas Health Observatory Indicator Booklets.

| Municipio | Altitude [masl] | T Average [$^{\circ}C$] | Municipio | Altitude [masl] | T Average [$^{\circ}C$] | Municipio | Altitude [masl] | T Average [$^{\circ}C$] |
| --- | --- | --- | --- | --- | --- | --- | --- | --- |
| Aguadas | 2214 | 18.5 | Manzanares | 1871 | 21.1 | Riosucio | 1380 | 20.2 |
| Anserma | 1720 | 19.4 | Marmato | 1370 | 22.1 | Risaralda | 1743 | 19 |
| Aranzazu | 1960 | 18 | Marquetalia | 1600 | 20.5 | Salamina | 1775 | 22 |
| Belalcazar | 1632 | 20.2 | Marulanda | 2825 | 13 | Samaná | 1460 | 19.8 |
| Chinchiná | 1380 | 20.3 | Neira | 1969 | 18 | San José | 1710 | 18 |
| Filadelfia | 1620 | 20 | Norcasia | 700 | 25 | Supía | 1183 | 21.6 |
| La Dorada | 178 | 31 | Pacora | 1819 | 18.4 | Victoria | 710 | 26 |
| La Merced | 1819 | 23 | Palestina | 1630 | 21.8 | Villamaría | 1920 | 18 |
| Manizales | 2150 | 19.9 | Pensilvania | 2100 | 19.7 | Viterbo | 998 | 24 |

**Table 3.** Estimated values of the model parameters.

| MunicipE | $\delta_h$ | $\beta$ | $\lambda_m$ | $\lambda_h$ | $\delta_m$ | $\mu$ | $\Lambda_h$ | $\Lambda_m$ min | $\Lambda_m$ max |
| --- | --- | --- | --- | --- | --- | --- | --- | --- | --- |
| Aguadas | $2,55\epsilon^{-5}$ | 0,17 | 0,44 | 0,45 | 0,04 | 0.329 | 0 | 0 | 0 |
| Anserma | $3\epsilon^{-5}$ | 0,18 | 0,51 | 0,52 | 0,04 | 0.329 | 36,7 | 30 | 300 |
| Aranzazu | $2,3\epsilon^{-5}$ | 0,17 | 0,41 | 0,41 | 0,03 | 0.329 | 0 | 0 | 0 |
| Belalcazar | $1,8\epsilon^{-5}$ | 0,18 | 0,57 | 0,58 | 0,04 | 0.329 | 19,45 | 13,66 | 147,36 |
| Chinchiná | $2,52\epsilon^{-5}$ | 0,18 | 0,58 | 0,59 | 0,04 | 0.329 | 50,88 | 48,63 | 533,08 |
| Filadelfia | $3,45\epsilon^{-5}$ | 0,18 | 0,55 | 0,57 | 0,04 | 0.329 | 7,45 | 2 | 83,3 |
| La Dorada | $1,58\epsilon^{-5}$ | 0,23 | 1,36 | 0,73 | 0,03 | 0.329 | 354,7 | 86,6 | 821,3 |
| La Merced | $3,67\epsilon^{-5}$ | 0,19 | 0,77 | 0,79 | 0,03 | 0.329 | 0 | 0 | 0 |
| Manizales | $2,79\epsilon^{-5}$ | 0,18 | 0,55 | 0,56 | 0,04 | 0.329 | 0 | 0 | 0 |
| Manzanares | $2,49\epsilon^{-5}$ | 0,18 | 0,63 | 0,65 | 0,04 | 0.329 | 0 | 0 | 0 |
| Marmato | $1,49\epsilon^{-5}$ | 0,19 | 0,71 | 0,73 | 0,04 | 0.329 | 19,25 | 15,88 | 277,58 |
| Marquetalia | $2,26\epsilon^{-5}$ | 0,18 | 0,59 | 0,61 | 0,04 | 0.329 | 48,9 | 33,9 | 339 |
| Marulanda | $3,34\epsilon^{-5}$ | 0,15 | 0,04 | 0,04 | 0,05 | 0.329 | 0 | 0 | 0 |
| Neira | $2,92\epsilon^{-5}$ | 0,17 | 0,41 | 0,41 | 0,03 | 0.329 | 29,66 | 3,65 | 40,72 |
| Norcasia | $2,29\epsilon^{-5}$ | 0,2 | 0,92 | 0,91 | 0,03 | 0.329 | 7,16 | 6,32 | 112,71 |
| Pacora | $2,84\epsilon^{-5}$ | 0,17 | 0,44 | 0,44 | 0,04 | 0.329 | 0 | 0 | 0 |
| Palestina | $4,50\epsilon^{-5}$ | 0,19 | 0,69 | 0,71 | 0,04 | 0.329 | 19,872 | 9,3 | 497 |
| Pensilvania | $2,85\epsilon^{-5}$ | 0,18 | 0,53 | 0,54 | 0,04 | 0.329 | 3,46 | 1,65 | 36,5 |
| Riosucio | $2,64\epsilon^{-5}$ | 0,18 | 0,57 | 0,58 | 0,04 | 0.329 | 13,94 | 3,94 | 139,38 |
| Risaralda | $2,67\epsilon^{-5}$ | 0,18 | 0,48 | 0,49 | 0,04 | 0.329 | 0 | 0 | 0 |
| Salamina | $3,37\epsilon^{-5}$ | 0,19 | 0,70 | 0,72 | 0,04 | 0.329 | 0 | 0 | 0 |
| Samaná | $2,76\epsilon^{-5}$ | 0,18 | 0,54 | 0,55 | 0,04 | 0.329 | 16,9 | 6,9 | 169 |
| San José | $2,624\epsilon^{-5}$ | 0,17 | 0,41 | 0,41 | 0,03 | 0.329 | 14,41 | 4,41 | 44,1 |
| Supía | $2,07\epsilon^{-5}$ | 0,187 | 0,67 | 0,69 | 0,04 | 0.329 | 16,03 | 5,64 | 204,57 |
| Victoria | $2,66\epsilon^{-5}$ | 0,21 | 0,99 | 0,95 | 0,03 | 0.329 | 5,69 | 4,39 | 44 |
| Villamaría | $2,24\epsilon^{-5}$ | 0,17 | 0,41 | 0,41 | 0,03 | 0,32 | 6,58 | 3,65 | 36,5 |
| Viterbo | $2,52\epsilon^{-5}$ | 0,19 | 0,85 | 0,85 | 0,03 | 0,35 | 23,73 | 15,71 | 308,05 |

With the demographic, epidemiological and mobility parameters, initial conditions were set for each municipality (presented in Table 4). To calibrate the model to capture the number of dengue cases historically occurred in the department of Caldas (CO) during some outbreaks in the last decade, periodic perturbations in the number of infected mosquitoes during two rainy seasons in a year were introduced. As a necessary simplification, we only considered one dengue serotype; this assumption implies that individuals can only be infected once. The model was adjusted by minimizing the error between simulated and real cases. In particular, the SIR-SI simulation model with the 33 municipalities connected after calibration yields 1099 cases of dengue in Caldas (see Table 6).

**Table 4.** Model initial conditions

| Municipe | $S_m(0)$ | $I_{mRain}(0)$ | $I_{mDrynees}(0)$ | $S_h(0)$ | $I_h(0)$ | $R_h(0)$ |
| --- | --- | --- | --- | --- | --- | --- |
| Aguadas | 0 | 0 | 0,001 | 22081 | 0 | 0 |
| Anserma | 37,704 | 4,189 | 0,001 | 33792 | 0 | 0 |
| Aranzazu | 0 | 0 | 0,001 | 11422 | 0 | 0 |
| Belalcazar | 17,167 | 1,907 | 0,001 | 10863 | 0 | 0 |
| Chinchiná | 38,763 | 4,307 | 0,001 | 51492 | 0 | 0 |
| Filadelfía | 5,553 | 0,617 | 0,001 | 11034 | 0 | 0 |
| La Dorada | 57,411 | 6,379 | 0,001 | 76963 | 0 | 0 |
| La Merced | 0 | 0 | 0,001 | 5508 | 0 | 0 |
| Manizales | 0 | 0 | 0,001 | 396075 | 0 | 0 |
| Manzanares | 0 | 0 | 0,001 | 23274 | 0 | 0 |
| Marmato | 165,579 | 18,398 | 0,001 | 9096 | 0 | 0 |
| Marquetalia | 18,520 | 2,058 | 0,001 | 14992 | 0 | 0 |
| Marulanda | 0 | 0 | 0,001 | 3406 | 0 | 0 |
| Neira | 3,713 | 0,413 | 0,001 | 30513 | 0 | 0 |
| Norcasia | 62,603 | 6,956 | 0,001 | 6374 | 0 | 0 |
| Pácora | 0 | 0 | 0,001 | 11952 | 0 | 0 |
| Palestina | 79,036 | 8,782 | 0,001 | 17760 | 0 | 0 |
| Pensilvania | 50,405 | 5,601 | 0,001 | 26361 | 0 | 0 |
| Riosucio | 34,613 | 3,846 | 0,001 | 61535 | 0 | 0 |
| Risaralda | 0 | 0 | 0,001 | 9583 | 0 | 0 |
| Salamina | 0 | 0 | 0,001 | 16635 | 0 | 0 |
| Samaná | 63,076 | 7,008 | 0,001 | 25777 | 0 | 0 |
| San José | 5,549 | 0,617 | 0,001 | 7588 | 0 | 0 |
| Supía | 14,036 | 1,560 | 0,001 | 26728 | 0 | 0 |
| Victoria | 5,253 | 0,584 | 0,001 | 8415 | 0 | 0 |
| Villamaría | 9,283 | 1,031 | 0,001 | 56303 | 0 | 0 |
| Viterbo | 10,244 | 1,138 | 0,001 | 12469 | 0 | 0 |

**Table 5.**
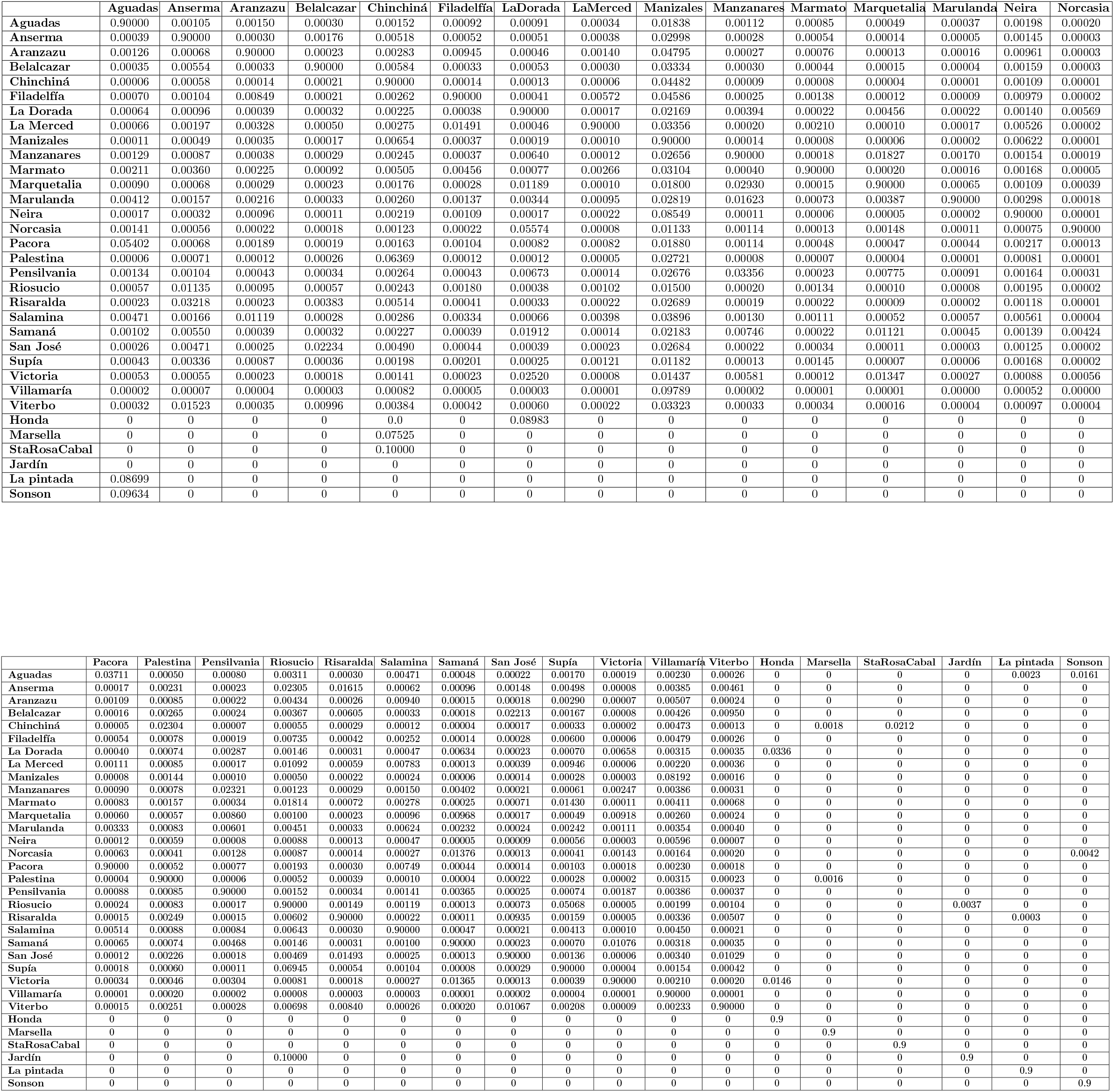
Matrix ϒ. Each cell represents the probability that an inhabitant of a municipality moves to another one.

**Table 6.** Dengue cases full conection

| Municipio | # cases | Municipio | # cases | Municipio | # cases |
| --- | --- | --- | --- | --- | --- |
| Aguadas | 1 | Manzanares | 2 | Riosucio | 41 |
| Anserma | 16 | Marmato | 204.8 | Risaralda | 0.4 |
| Aranzazu | 0.4 | Marquetalia | 14.2 | Salamina | 1.4 |
| Belalcazar | 9.4 | Marulanda | 0.0 | Samaná | 41.8 |
| Chinchiná | 128.2 | Neira | 2.8 | San José | 3.1 |
| Filadelfia | 4.5 | Norcasia | 129.9 | Supía | 46.7 |
| La Dorada | 481.0 | Pacora | 0.0 | Victoria | 13.4 |
| La Merced | 0.4 | Palestina | 47.3 | Villamaría | 9.4 |
| Manizales | 312.0 | Pensilvania | 25.2 | Viterbo | 22.4 |
| Total cases Caldas | 1099 |  |  |  |  |

**Table 7.** Dengue incidence rate per 100 000 inhabitants, by municipality of origin. Source: Public Health Observatory.

| **Municipio** | **2015** | **2016** | **2017** | **2018** | **2019** | **2020** | **Total municipio** |
| --- | --- | --- | --- | --- | --- | --- | --- |
| Marmato | 2836,4 | 571,7 | 120,9 | 55,0 | 77,0 | 22,0 | 3682,9 |
| Norcasia | 2682,8 | 47,1 | 0,0 | 31,4 | 627,5 | 31,4 | 3420,1 |
| Viterbo | 248,6 | 200,5 | 24,1 | 0,0 | 312,8 | 24,1 | 810,0 |
| Risaralda | 20,9 | 427,8 | 41,7 | 20,9 | 0,0 | 167,0 | 678,3 |
| La Dorada | 157,2 | 46,8 | 7,8 | 18,2 | 283,3 | 52,0 | 565,2 |
| Supía | 138,4 | 396,6 | 7,5 | 3,7 | 7,5 | 3,7 | 557,5 |
| Palestina | 264,6 | 163,3 | 28,2 | 5,6 | 39,4 | 28,2 | 529,3 |
| Chinchiná | 112,6 | 324,3 | 33,0 | 5,8 | 21,4 | 25,2 | 522,4 |
| Victoria | 83,2 | 11,9 | 11,9 | 23,8 | 118,8 | 154,5 | 404,0 |
| Marquetalia | 73,4 | 186,8 | 33,4 | 0,0 | 66,7 | 13,3 | 373,5 |
| Filadelfia | 9,1 | 308,1 | 0,0 | 0,0 | 0,0 | 0,0 | 317,2 |
| Samaná | 120,3 | 42,7 | 3,9 | 11,6 | 42,7 | 7,8 | 228,9 |
| Anserma | 50,3 | 115,4 | 3,0 | 0,0 | 0,0 | 8,9 | 177,6 |
| Riosucio | 37,4 | 134,9 | 1,6 | 0,0 | 1,6 | 1,6 | 177,1 |
| Aguadas | 122,3 | 13,6 | 27,2 | 0,0 | 0,0 | 0,0 | 163,0 |
| Belalcazar | 73,6 | 46,0 | 0,0 | 0,0 | 0,0 | 9,2 | 128,9 |
| Pensilvania | 98,6 | 19,0 | 0,0 | 3,8 | 0,0 | 0,0 | 121,4 |
| Salamina | 18,0 | 72,1 | 6,0 | 0,0 | 0,0 | 6,0 | 102,2 |
| La Merced | 36,3 | 36,3 | 0,0 | 0,0 | 0,0 | 0,0 | 72,6 |
| Manizales | 38,1 | 2,3 | 3,0 | 0,3 | 0,0 | 1,5 | 45,2 |
| Villamaría | 7,1 | 14,2 | 3,6 | 0,0 | 1,8 | 0,0 | 26,6 |
| San José | 13,2 | 13,2 | 0,0 | 0,0 | 0,0 | 0,0 | 26,4 |
| Neira | 3,3 | 3,3 | 3,3 | 0,0 | 0,0 | 3,3 | 13,1 |
| Pácora | 8,4 | 0,0 | 0,0 | 0,0 | 0,0 | 0,0 | 8,4 |
| Manzanares | 0,0 | 0,0 | 0,0 | 0,0 | 4,3 | 0,0 | 4,3 |
| Aranzazu | 0,0 | 0,0 | 0,0 | 0,0 | 0,0 | 0,0 | 0,0 |
| Marulanda | 0,0 | 0,0 | 0,0 | 0,0 | 0,0 | 0,0 | 0,0 |
| **Total** | **7254,1** | **3197,8** | **359,9** | **180,1** | **1604,7** | **559,6** |  |

The agreement between real cases and those obtained after the SIR-SI model has been calibrated is shown in Fig. 6. In particular the map on the left shows the dengue cases reported by the health entities and on the right the dengue cases estimated with the model. It can be seen at the bottom that there is a correlation of 0.76 between the data estimated with the model and the cases occurred in the department of Caldas during 2015.

**Fig 6.**
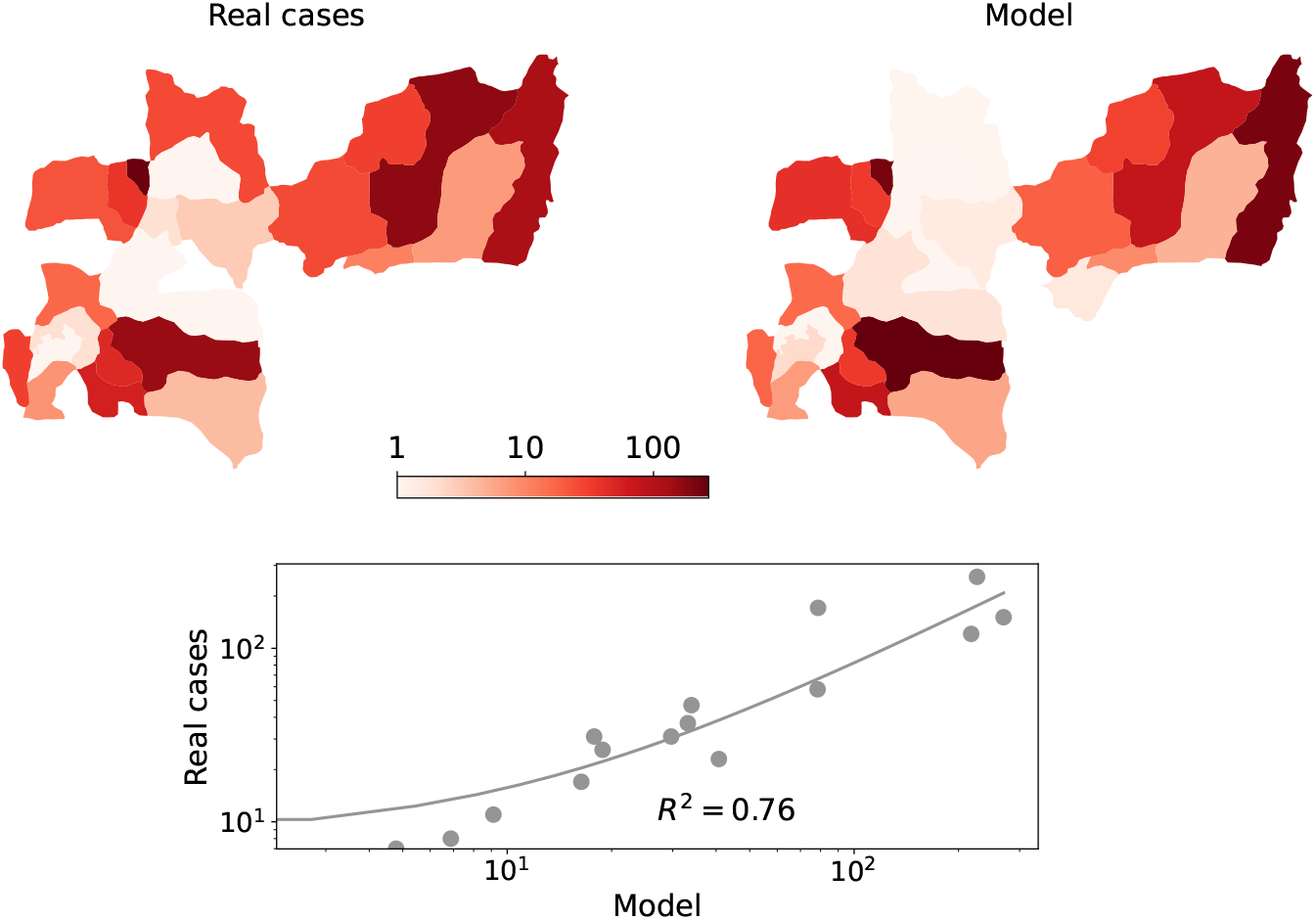
On the left, cases of dengue fever occurred in the department of Caldas during one year (2015). On the right, dengue cases predicted by the model during the same year. Below correlation between both variables.

### 3.3 Control Scenario 1: Mobility restrictions focused on high incidence nodes

The purpose with this scenario is to get some insights on the contribution to the total number of dengue cases in the network, after applying mobility constraints to the five most affected cities (one at a time). We analyze the variation of dengue cases according to mobility constraints when: i) Access: No foreigners are allowed to enter the municipality, ii) Exit: no inhabitants can leave the municipality, iii) Isolation: No entry or exit allowed for residents or visitors. These control measures were applied in simulations for the five municipalities with the highest number of dengue cases during during the period 2015 − 2019 according to the public health office of Caldas [46]. These municipalities are La Dorada, Norcasia, Marmato, Chinchiná and Manizales. Fig. 7 shows the variation of the total number of dengue cases in the department of Caldas when the three mobility control measures proposed in this work are applied in the five municipalities mentioned. In Fig.7 it can be seen that the total number of dengue cases in the department varies as mobility in each municipality is reduced.

**Fig 7.**
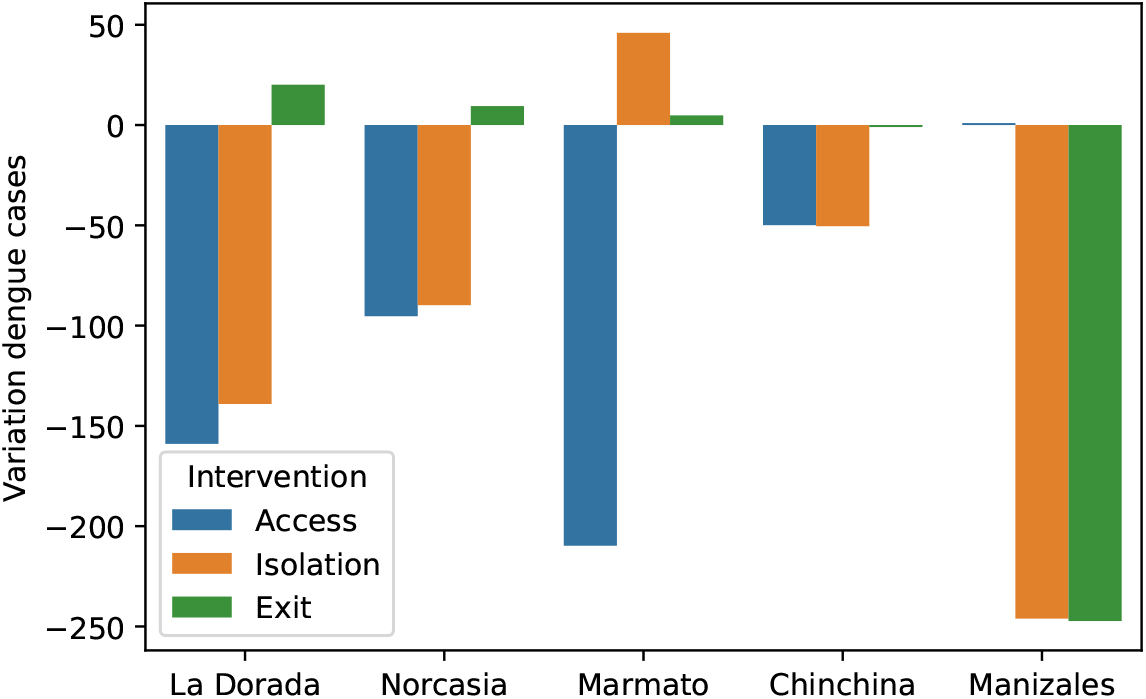
Variation of dengue cases in the department Caldas according to mobility restrictions when: i) no foreigners are allowed to enter the municipality (blue), no inhabitants can leave the municipality (green), the municipality is totally isolated (orange).

#### 3.3.1 Access

This control measure, when applied in each of the five municipalities with the highest burden of the disease, reduces the total number of dengue cases in the department. In Fig 7 it can be seen that the most significant reduction occurs when this control measure is applied in La Dorada and Marmato, with a decrease of 159 and 210, respectively. These values indicate that completely restricting access to this endemic municipality has a positive effect on reducing the total number of dengue cases in the department.

#### 3.3.2 Exit

When the exit of the inhabitants of the municipality of La Dorada is restricted, but the entry of foreigners is allowed, the number of dengue cases increase in 20 with respect to the free mobility scenario. This same behavior occurs when the control measure is applied in Norcasia and Marmato where the increase of 9 and 5 dengue cases. When the control measure is applied in Manizales, dengue cases are reduced by 247. This is due to the fact that its inhabitants are infected when traveling to endemic areas.

#### 3.3.3 Isolation

Isolating a municipality in this study means limiting mobility to zero, therefore its inhabitants cannot travel to other municipalities and people are not allowed to enter. Applying this control measure in La Dorada, Norcasia, Chinchiná and Manizales reduces the total number of dengue cases in the department by 139, 90, 51 and 270, respectively. The opposite occurs when applied in Marmato, where dengue cases increase by 46.

### 3.4 Control scenario 2: Confinement

During the COVID-19 pandemic, governmental entities with the objective of containing the spread of the virus have opted to isolate neighborhoods, even entire countries. In this work we simulate the effect of confining the municipalities in order to analyze how this restriction influences the total number of dengue cases. Confinement consists of limiting the exit of inhabitants and entry of foreigners to each municipality. When confinement is applied, 630 cases occur, which implies a reduction of 42.67% with respect to the 1099 dengue cases that occurred when there are no restrictions. The dengue cases before and after confinement are shown in Table 8, where it can be seen that mobility affects each municipality differently. The column (variation of cases) shows positive values when the mobility restriction reduces dengue cases and negative values when there is an inverse effect. It can be observed that in the municipalities of Manzanares, Salamina, Aguadas, Risaralda, Aranzazu, La Merced and Pácora, confining population reduces dengue cases to zero. Confinement reduces dengue cases in the municipalities of La Dorada, Victoria and Riosucio, where a higher percentage of travelers go to areas with a higher incidence of dengue. A similar phenomenon occurs in Norcasia, Marmato and Palestina, but in the opposite direction, with the highest percentage of travelers from these three municipalities going to areas with lower incidence. The percentages of travelers to municipalities of zero, lower and higher incidence than the municipality of residence can be seen in Table 9. The results obtained in this scenario coincide with those obtained by *Conceição* in his study conducted in Sao Paulo [47] where there was a reduction in dengue cases as a collateral result of population confinement to hinder the advance of COVID-19 pandemic.

**Table 8.** Dengue cases with and without mobility restrictions. Negative number means increase and positive number represents reduce after applying the mobility control measure (quarantine).

| Municipe | Unrestricted mobility | Restricted mobility | Variation in dengue cases | % of variation |
| --- | --- | --- | --- | --- |
| Manizales | 270 | 0 | +270 | 270.0 |
| La Dorada | 217 | 72 | +145 | 201.3 |
| Chinchina | 78 | 27 | +51 | 185.5 |
| Riosucio | 41 | 13 | +28 | 213.3 |
| Supia | 33 | 15 | +18 | 115.3 |
| Villamaria | 6 | 1 | +5 | 566.1 |
| Anserma | 16 | 14 | +2 | 13.0 |
| Victoria | 5 | 3 | +2 | 51.4 |
| Manzanares | 2 | 0 | +2 | 2.0 |
| Salamina | 1 | 0 | +1 | 1.0 |
| Filadelfia | 2 | 1 | +1 | 144.6 |
| Neira | 2 | 1 | +1 | 141.3 |
| Aguadas | 1 | 0 | +1 | 1.0 |
| La Merced | 0 | 0 | 0 | 0 |
| Aranzazu | 0 | 0 | 0 | 0 |
| Samana | 30 | 29 | +1 | 1 |
| Risaralda | 0 | 0 | 0 | 0.0 |
| Pacora | 0 | 0 | 0 | 0.0 |
| San Jose | 2 | 2 | 0 | 0 |
| Marulanda | 0 | 0 | 0 | 0.0 |
| Marquetalia | 9 | 9 | 0 | 0 |
| Belalcazar | 7 | 7 | 0 | 0 |
| Pensilvania | 19 | 20 | -1 | 6.5 |
| Palestina | 34 | 36 | -2 | 4.9 |
| Viterbo | 18 | 20 | -2 | 10.4 |
| Norcasia | 79 | 97 | -18 | 18.5 |
| Marmato | 226 | 261 | -35 | 13.5 |
| **Total** | **1099** | **630** | **+469** | **74.5** |

**Table 9.** Percentage of travelers to 0, lower and higher incidence areas than their city of residence.

| Municipio | Incidence when mobility is restricted | % of population traveling to municipalities of incidence 0 | % of population traveling to municipality lower incidence | % of population traveling to municipality higher incidence |
| --- | --- | --- | --- | --- |
| Norcasia | 4687 | 1,53 | 8,04 | 0 |
| Marmato | 4147 | 4,30 | 5,70 | 0,0047 |
| Viterbo | 395 | 4,33 | 5,63 | 0,0372 |
| Palestina | 343 | 2,81 | 7 | 0,0314 |
| La Dorada | 246 | 2,82 | 3,12 | 4,0575 |
| Samaná | 176 | 3,32 | 4,21 | 2,4671 |
| Supía | 149 | 1,63 | 8,09 | 0,2824 |
| Chinchiná | 146 | 4,56 | 0,76 | 2,3761 |
| Victoria | 145 | 2,21 | 2,14 | 5,6534 |
| Marquetalia | 118 | 5,10 | 1,47 | 3,4342 |
| Pensilvania | 112 | 6,58 | 0,91 | 2,5139 |
| Belalcazar | 105 | 4,12 | 3,75 | 2,1300 |
| Anserma | 71 | 62,98 | 3,03 | 2,1326 |
| San José | 45 | 4,31 | 0,98 | 4,7087 |
| Riosucio | 40 | 2,07 | 0,57 | 6,9826 |
| Filadelfia | 38 | 6,46 | 1,46 | 2,0850 |
| Villamaría | 8 | 9,81 | 0,05 | 0,1417 |
| Neira | 6 | 8,77 | 0,00 | 1,2300 |

**Table 10.**
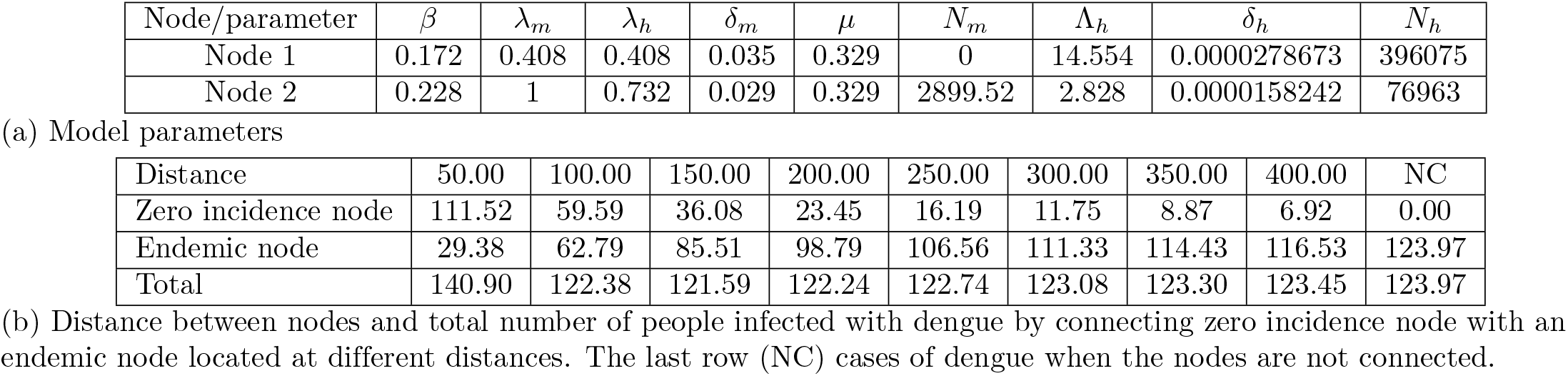
Example A.

| Node/parameter | $\beta$ | $\lambda_m$ | $\lambda_h$ | $\delta_m$ | $\mu$ | $N_m$ | $\Lambda_h$ | $\delta_h$ | $N_h$ |
| --- | --- | --- | --- | --- | --- | --- | --- | --- | --- |
| Node 1 | 0.172 | 0.408 | 0.408 | 0.035 | 0.329 | 0 | 14.554 | 0.0000278673 | 396075 |
| Node 2 | 0.228 | 1 | 0.732 | 0.029 | 0.329 | 2899.52 | 2.828 | 0.0000158242 | 76963 |
| (a) Model parameters |  |  |  |  |  |  |  |  |  |
| Distance | 50.00 | 100.00 | 150.00 | 200.00 | 250.00 | 300.00 | 350.00 | 400.00 | NC |
| Zero incidence node | 111.52 | 59.59 | 36.08 | 23.45 | 16.19 | 11.75 | 8.87 | 6.92 | 0.00 |
| Endemic node | 29.38 | 62.79 | 85.51 | 98.79 | 106.56 | 111.33 | 114.43 | 116.53 | 123.97 |
| Total | 140.90 | 122.38 | 121.59 | 122.24 | 122.74 | 123.08 | 123.30 | 123.45 | 123.97 |

(b) Distance between nodes and total number of people infected with dengue by connecting zero incidence node with an endemic node located at different distances. The last row (NC) cases of dengue when the nodes are not connected.

**Table 11.**
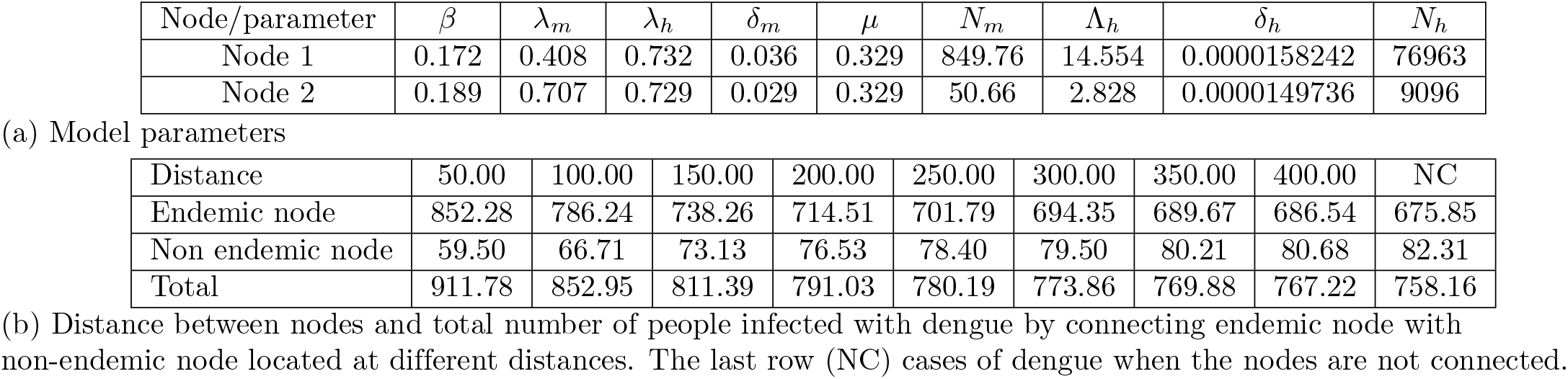
Example B.

| Node/parameter | $\beta$ | $\lambda_m$ | $\lambda_h$ | $\delta_m$ | $\mu$ | $N_m$ | $\Lambda_h$ | $\delta_h$ | $N_h$ |
| --- | --- | --- | --- | --- | --- | --- | --- | --- | --- |
| Node 1 | 0.172 | 0.408 | 0.732 | 0.036 | 0.329 | 849.76 | 14.554 | 0.0000158242 | 76963 |
| Node 2 | 0.189 | 0.707 | 0.729 | 0.029 | 0.329 | 50.66 | 2.828 | 0.0000149736 | 9096 |
| (a) Model parameters |  |  |  |  |  |  |  |  |  |
| Distance | 50.00 | 100.00 | 150.00 | 200.00 | 250.00 | 300.00 | 350.00 | 400.00 | NC |
| Endemic node | 852.28 | 786.24 | 738.26 | 714.51 | 701.79 | 694.35 | 689.67 | 686.54 | 675.85 |
| Non endemic node | 59.50 | 66.71 | 73.13 | 76.53 | 78.40 | 79.50 | 80.21 | 80.68 | 82.31 |
| Total | 911.78 | 852.95 | 811.39 | 791.03 | 780.19 | 773.86 | 769.88 | 767.22 | 758.16 |

(b) Distance between nodes and total number of people infected with dengue by connecting endemic node with non-endemic node located at different distances. The last row (NC) cases of dengue when the nodes are not connected.

**Table 12.**
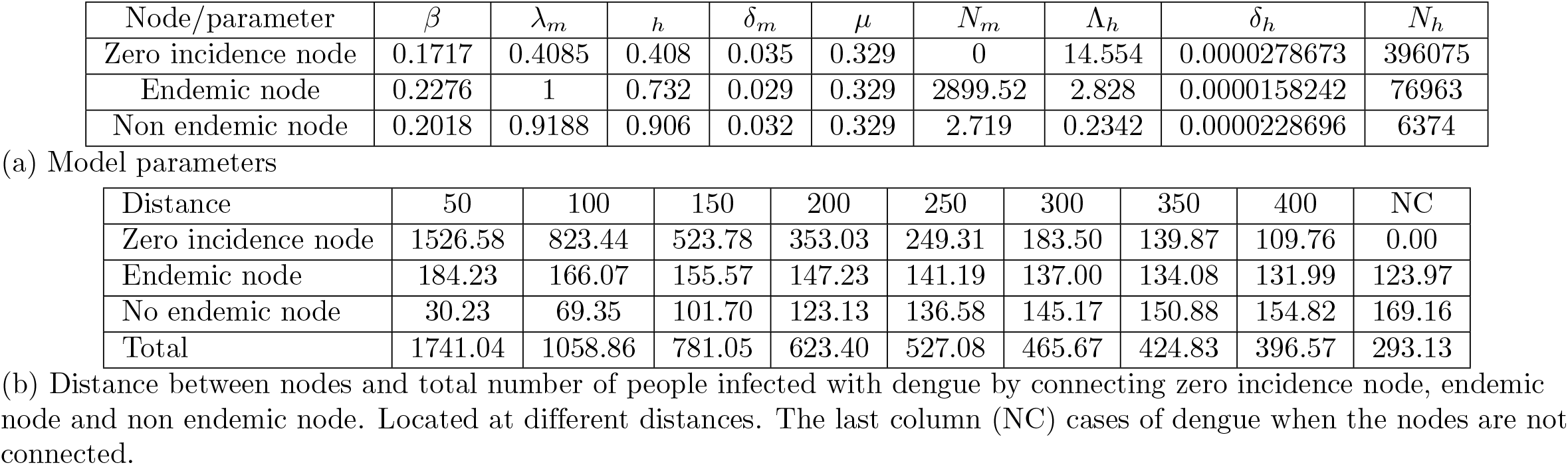
Example C.

| Node/parameter | $\beta$ | $\lambda_m$ | $h$ | $\delta_m$ | $\mu$ | $N_m$ | $\Lambda_h$ | $\delta_h$ | $N_h$ |
| --- | --- | --- | --- | --- | --- | --- | --- | --- | --- |
| Zero incidence node | 0.1717 | 0.4085 | 0.408 | 0.035 | 0.329 | 0 | 14.554 | 0.0000278673 | 396075 |
| Endemic node | 0.2276 | 1 | 0.732 | 0.029 | 0.329 | 2899.52 | 2.828 | 0.0000158242 | 76963 |
| Non endemic node | 0.2018 | 0.9188 | 0.906 | 0.032 | 0.329 | 2.719 | 0.2342 | 0.0000228696 | 6374 |

(a) Model parameters
| Distance | 50 | 100 | 150 | 200 | 250 | 300 | 350 | 400 | NC |
| --- | --- | --- | --- | --- | --- | --- | --- | --- | --- |
| Zero incidence node | 1526.58 | 823.44 | 523.78 | 353.03 | 249.31 | 183.50 | 139.87 | 109.76 | 0.00 |
| Endemic node | 184.23 | 166.07 | 155.57 | 147.23 | 141.19 | 137.00 | 134.08 | 131.99 | 123.97 |
| No endemic node | 30.23 | 69.35 | 101.70 | 123.13 | 136.58 | 145.17 | 150.88 | 154.82 | 169.16 |
| Total | 1741.04 | 1058.86 | 781.05 | 623.40 | 527.08 | 465.67 | 424.83 | 396.57 | 293.13 |

(b) Distance between nodes and total number of people infected with dengue by connecting zero incidence node, endemic node and non endemic node. Located at different distances. The last column (NC) cases of dengue when the nodes are not connected.

## 4 Discussion

Several scenarios were analyzed to provide policy makers with insight into the implementation of control measures to mitigate the spread of dengue in the department of Caldas. The study used a compartmental model at each node of a network and investigated the effects on the total number of dengue cases when applying mobility control measures such as Access: No foreigners are allowed to enter the municipality, Isolation: No residents or visitors are allowed to enter or leave, and Exit: No inhabitant is allowed to leave the municipality. The results obtained explain phenomena that have been observed in the historical data, and the coexistence of different types of trends according to local conditions and connectivity. Such is the case of the capital Manizales (zero incidence since environmental conditions are not suitable for the mosquito), which has reported cases of the disease in the health system. The inhabitants of this municipality contract the disease when moving to areas where dengue fever is present. The simulations show this phenomenon and it is observed that limiting mobility from nodes of zero incidence to nodes with the presence of the disease prevents the spread of the disease. The results obtained in this study are similar to those obtained by Jassen, who showed that bans on international air travel during the COVID-19 pandemic led to a sharp decrease in dengue cases in Australia, where the disease is predominantly imported [48]. Another phenomenon observed was that the isolation of Marmato (a node with a high incidence of the disease) generates an increase in dengue cases. Given that when human movements are limited to one’s own home and its surroundings, contact between people and vectors may increase, resulting in a higher risk of exposure and transmission of the virus, as occurred in Thailand where the isolation generated by COVID-19 generated an increase in dengue cases [49]. On the contrary, when residents of Marmato travel to nearby municipalities with a lower incidence of the disease, the probability of contagion is reduced with displacement. In the case of La Dorada, which is also a municipality with a high incidence, the increase in cases is generated by the departure of its inhabitants. This is due to the fact that most people move to areas with a higher incidence of the disease, which increases the probability of infection and, therefore, the number of infected people. This phenomenon was evidenced in Singapore where the mobility of inhabitants was limited to public spaces, such as workplaces or schools, where the density of mosquitoes was high, which led to a decrease in dengue transmission during the COVID-19 quarantine [50]. Therefore, in La Dorada or any municipality with these mobility characteristics, the control measure that generates the greatest impact on the reduction of dengue cases is to limit the exit of its inhabitants. In this study it was observed that the reduction of dengue cases after applying quarantine is not significantly higher than applying other measures, which is why it is suggested to control entities analyze the relevance of this measure when making decisions since, as has been observed in the current pandemic, isolation has a negative influence on people. It causes alterations in eating habits [51], increases the use of drugs [52], has negative psychological [53, 54], and economic [55] effects on people around the world. The analysis carried out in this study can be used, both in the department of Caldas as well as in any other region, to design mobility reduction strategies to control or mitigate a dengue outbreak.

## 5 Conclusion

The model presented in this study makes it possible to analyze the effect of applying control measures to reduce dengue cases in a region. By comparing the results of the model with the actual cases, it has been established that the estimated values have a realistic correspondence. Based on the results, it is recommended that health experts and administrative authorities limit mobility to areas with high incidence of the disease during dengue outbreaks in order to prevent its spread. This model can be extrapolated to any region, if the average temperature, altitude, number of inhabitants, human vital statistics and aedic records are known. Moreover, the mobility control measures here discussed can be taken into account for the development of public health policies in endemic regions.

## Data Availability

All relevant data are within the manuscript and its Supporting Information files.

## 6 Acknowledgments

We acknowledge the Health observatory of Caldas, Territorial of Health of Caldas and National Coffee Research Center - Cenicafé for allowing access to the database and departmental information for the development of this study. Carolina Ospina acknowledges funding from Colciencias “convocatoria doctorados nacionales 647 de 2014”. We acknowledge financial support from grant PID2020-113582GB-I00 funded by MCIN/AEI/10.13039/501100011033, and from Departamento de Industria e Innovación del Gobierno de Aragón y Fondo Social Europeo through projects no. E36 17R (FENOL group).

## 7 Supporting information

